# Absent or insufficient anti-SARS-CoV-2 S antibodies at ICU admission are associated to higher viral loads in plasma, antigenemia and mortality in COVID-19 patients

**DOI:** 10.1101/2021.03.08.21253121

**Authors:** María Martin-Vicente, Raquel Almansa, Isidoro Martínez, Ana P. Tedim, Elena Bustamante, Luis Tamayo, César Aldecoa, José Manuel Gómez, Gloria Renedo, Jose Ángel Berezo, Jamil Antonio Cedeño, Nuria Mamolar, Pablo García Olivares, Rubén Herrán, Ramón Cicuendez, Pedro Enríquez, Alicia Ortega, Noelia Jorge, Amanda de la Fuente, Juan Bustamante-Munguira, María José Muñoz-Gómez, Milagros González-Rivera, Carolina Puertas, Vicente Más, Mónica Vázquez, Felipe Pérez-García, Jesús Rico-Feijoo, Silvia Martín, Anna Motos, Laia Fernandez-Barat, Jose María Eiros, Marta Dominguez-Gil, Ricard Ferrer, Ferrán Barbé, David J Kelvin, Jesús F Bermejo-Martin, Salvador Resino, Antoni Torres

## Abstract

**Purpose:** to evaluate the association between anti-SARS-CoV-2 S IgM and IgG antibodies with viral RNA load in plasma, the frequency of antigenemia and with the risk of mortality in critically ill patients with COVID-19.

**Methods:** anti-SARS-CoV-2 S antibodies levels, viral RNA load and antigenemia were profiled in plasma of 92 adult patients in the first 24 hours following ICU admission. The impact of these variables on 30-day mortality was assessed by using Kaplan-Meier curves and multivariate Cox regression analysis.

**Results:** non survivors showed more frequently absence of anti-SARS-CoV-2 S IgG and IgM antibodies than survivors (26.3% vs 5.6% for IgM and 18.4% vs 5.6% for IgG), and a higher frequency of antigenemia (47.4% vs 22.2%) (*p* <0.05). Non survivors showed lower concentrations of anti-S IgG and IgM and higher viral RNA loads in plasma, which were associated to increased 30-day mortality and decreased survival mean time. [Adjusted HR (CI95%), *p*]: [S IgM (AUC ≥60): 0.48 (0.24; 0.97), 0.040]; [S IgG (AUC ≥237): 0.47 (0.23; 0.97), 0.042]; [Antigenemia (+): 2.45 (1.27; 4.71), 0.007]; [N1 viral load (≥ 2.156 copies/mL): 2.21 (1.11; 4.39),0.024]; [N2 viral load (≥ 3.035 copies/mL): 2.32 (1.16; 4.63), 0.017]. Frequency of antigenemia was >2.5-fold higher in patients with absence of antibodies. Levels of anti-SARS-CoV-2 S antibodies correlated inversely with viral RNA load.

**Conclusion:** absence / insufficient levels of anti-SARS-CoV-2 S antibodies following ICU admission is associated to poor viral control, evidenced by increased viral RNA loads in plasma, higher frequency of antigenemia, and also to increased 30-day mortality.

**Take-home message:** absent or low levels of antibodies against the S protein of SARS-CoV- 2 at ICU admission is associated to an increased risk of mortality, higher frequency of antigenemia and higher viral RNA loads in plasma. Profiling anti-SARS-CoV-2 s antibodies at ICU admission could help to predict outcome and to better identify those patients potentially deserving replacement treatment with monoclonal or polyclonal antibodies.

## Introduction

Anti-SARS-CoV-2 S antibodies bind to the viral spike protein, inhibiting virus attachment to cell surface receptors [1]. Therefore, during infection with SARS-CoV-2, the development of anti-S antibodies could reduce viral replication by interfering with virus entry into a cell. From the clinical point of view, recent evidence suggests that anti-SARS-CoV-2 S antibodies could play a role in protecting against severe disease in patients with COVID-19 [2]. Nonetheless, the impact of these antibodies on patients’ survival with COVID-19 admitted to the ICU has not been sufficiently addressed to the present moment.

Recent works from our group and others have evidenced the importance of SARS-CoV-2 RNAemia [3] [4] [5] and antigenemia as markers of severity in COVID-19 [6]. The presence of viral material in plasma could represent a surrogated marker of poor viral control by patient’s immune response. Whether anti-SARS-CoV-2 S antibodies could have any influence on the dissemination of viral genomic material or viral proteins at the systemic level has yet to be properly studied.

In this work, we profiled levels of anti-SARS-CoV-2 S IgM and IgG antibodies in plasma of 92 patients with COVID-19 in the first 24 hours following admission to the ICU, and evaluated their association with mortality. In parallel, we quantified viral RNA load in plasma and tested the presence or absence of antigenemia in these patients, correlating them with the levels of antibodies and outcome.

## Materials and Methods

### Study design

92 critically ill adult patients with a positive nasopharyngeal swab polymerase chain reaction (PCR) test for SARS-CoV-2 performed at participating hospitals were recruited during the first pandemic wave in Spain from March 16^th^ to April 15^th^ 2020.

### Blood samples

Plasma from blood collected in EDTA tubes samples was obtained in the first 24 hours following admission to the ICU, following proper centrifugation. Samples were stored at −80°C until quantification of antibodies, viral RNA load and antigenemia evaluation.

### Detection and quantification of SARS-CoV-2 RNA in plasma

RNA was extracted from 100 µl of plasma using an automated system, eMAG® from bioMérieux® (Marcy l’Etoile, France). Detection and quantification of SARS-CoV-2 RNA was performed in five µl of the eluted solution using the Bio-Rad SARS-CoV-2 ddPCR kit according to manufacturer’s specifications on a QX-200 droplet digital PCR platform from the same provider. This PCR targets the N1 and N2 regions of the viral nucleoprotein gene and also the human ribonuclease (RNase) P gene using the primers and probes sets detailed in the CDC 2019-Novel Coronavirus (2019-nCoV) Real-Time RT-PCR Diagnostic Panel [7]. Samples were considered positive for SARS-CoV-2 when N1 and/or N2 presented values ≥ 0.1 copies/µL in a given reaction. RNase P gene was considered positive when it presented values ≥ 0.2 copies/µL, following manufacturer’s indications. The test was only considered valid when RNase P gene was positive. Final results were given in copies of cDNA / mL of plasma.

### Immunoassay for antibody quantification

a specific immunoassay was developed to quantify anti-SARS-CoV-2 S IgG and IgM antibodies in plasma. The plasmid pαH coding for the S protein ectodomain (residues 1-1208) of the SARS-CoV-2 2019-nCOV (GenBank: MN908947) was kindly provided by Dr. Jason McLellan (the University of Texas at Austin- USA) [8]. Mutagenesis was carried out to obtain a HexaPro construct that allowed a high- yield production of a stabilized prefusion spike protein [9]. The following substitutions were included at the ectodomain: glycine at residue 614 (D614G), a “GSAS” substitution at the furin cleavage site (residues 682–685), and proline at residues 817, 892, 899, 942, 986, and 987. For trimerization and purification, the C-terminal end of the S protein ectodomain was fused to the T4 fibritin trimerization motif (foldon), an HRV3C protease cleavage site, and an 8XHisTag. The expression vector coding for the SARS-CoV-2 S protein ectodomain was used to transiently transfect FreeStyle 293F cells (Thermo Fisher, Waltham, MA, USA) using polyethylenimine. The S protein domain was purified from filtered cell supernatants using Ni-NTA resins (Sigma Aldrich, San Luis, MO, USA) and subjected to an additional purification step by size-exclusion chromatography using a Superose 6 10/300 column (GE Healthcare, Chicago, IL, USA).

### Antibody titration

Antibody titers against the S protein were determined by incubating serial dilutions of serum samples (starting at a 1:50 dilution) with the purified S protein ectodomain. Ninety-six well plates were coated with 200 ng per well of the S protein ectodomain. The following day, serum samples were added, and the binding to the S protein was determined by successive incubations with a secondary peroxidase-conjugated anti-human IgG or IgM (Jackson Immunoresearch, West Grove, PA, USA) antibody and the OPD substrate (Sigma Aldrich, San Luis, MO, USA). The area under the curve (AUC) was calculated by using GraphPad Prism 8.0 (GraphPad Sofware, Inc., San Diego, CA, USA) and the following parameters: Baseline Y=0.1; ignore peaks that are less than 10% of the distance from minimum to maximum Y; all peaks must go above the baseline. The AUC is expressed as X units times the Y units.

### Antigenemia

The presence/absence of N antigen of SARS-CoV-2 in plasma was evaluated by using the Panbio™ COVID-19 Ag Rapid Test Device from Abbott (Chicago, IL, USA).

### Outcome and factor variables

The outcome variable was mortality at 30 days. The factors analyzed were: i) plasma IgM titers against SARS-CoV-2 spike protein; ii) plasma IgG titers against SARS-CoV-2 spike protein; iii) plasma SARS-CoV-2 N-antigenemia; iv) plasma SARS-CoV-2 N1-RNA; v) plasma SARS-CoV-2 N2-RNA.

### Statistical analysis

Statistical analysis was performed using IBM SPSS Statistics 25.0 (SPSS INC, Armonk, NY, U.S.A), and figures were generated using GraphPad Prism 8.0 (GraphPad Software, Inc., San Diego, CA, U.S.A). The level of significance was set at 0.05 (2-tailed). For descriptive analysis of patient characteristics, the differences between independent groups were assessed using the Chi-square test or Fisher’s Exact Test for categorical variables. Differences for continuous variables were evaluated by using the Mann-Whitney U test. The ability of the factor variables to differentiate survivors from non-survivors was evaluated using the receiver-operating characteristic (ROC) curve. The cut-off of those factor variables for 30-day mortality prediction was obtained by calculating the optimal operating point (OOP) in the ROC, namely, the point on the ROC that had the minimum distance to the upper left corner calculated by Pythagorean theorem.

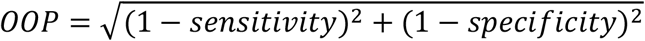

For association analysis, survival analysis was carried out to evaluate 30-day mortality. According to presence/absence and OOP for the ROC curve, these factor variables were stratified into categorical variables. Kaplan-Meier product-limit method was used to calculate survival probabilities and the log-rank test to compare groups. We also used Cox proportional-hazards models to estimate the risk of dying, adjusted by the significant covariates at baseline (see **Table 1**).

**Table 1:** Baseline characteristics of patients admitted to the intensive care unit. **Statistics:** Continuous variables are represented as median (interquartile range) and categorical variables as absolute count (%). *P-values* were calculated by Mann-Whitney for continuous variables and chi-squared tests or Fisher’s exact test for categorical variables. Significant differences are shown in bold. **Abbreviations:** p-value, level of significance; INR, international normalized ratio; COPD, chronic obstructive pulmonary disease; LDH, lactic acid dehydrogenase; GPT, glutamic-pyruvate transaminase; WBC, white blood cell; APACHE II, Acute Physiology and Chronic Health disease Classification System II; SOFA, Sequential Organ Failure Assessment, ARDS, acute respiratory distress syndrome.

| Characteristics | All | Alive by day 30 | Dead by day 30 | P-value |
| --- | --- | --- | --- | --- |
| **No.** | 92 | 54 | 38 |  |
| **Epidemiology** |  |  |  |  |
| **Age (years)** | 66 (50; 71.5) | 60 (47; 67) | 70 (66; 75) | **<0.001** |
| **Male** | 60 (65.2%) | 37 (68.5%) | 23 (60.5%) | 0.428 |
| **Alcoholism** | 1 (1.1%) | 0 (0%) | 1 (2.6%) | 0.413 |
| **Smoking** | 5 (5.4%) | 2 (3.7%) | 3 (7.9%) | 0.645 |
| **Comorbidities** |  |  |  |  |
| **Cardiac disease** | 7 (7.6%) | 2 (3.7%) | 5 (13.2%) | 0.121 |
| **Chronic vascular disease** | 5 (5.4%) | 4 (7.4%) | 1 (2.6%) | 0.401 |
| **COPD** | 3 (3.3%) | 2 (3.7%) | 1 (2.6%) | 0.999 |
| **Asthma** | 2 (2.2%) | 1 (1.9%) | 1 (2.6%) | 0.999 |
| **Obesity** | 22 (23.9%) | 16 (29.6%) | 6 (15.8%) | 0.125 |
| **Hypertension** | 39 (42.4%) | 17 (31.5%) | 22 (57.9%) | **0.012** |
| **Dyslipidemia** | 30 (32.6%) | 17 (31.5%) | 13 (34.2%) | 0.783 |
| **Chronic renal disease** | 3 (3.3%) | 1 (1.9%) | 2 (5.3%) | 0.567 |
| **Chronic hepatic disease** | 3 (3.3%) | 1 (1.9%) | 2 (5.3%) | 0.567 |
| **Type 1 diabetes** | 3 (3.3%) | 3 (5.6%) | 0 (0%) | 0.265 |
| **Type 2 diabetes** | 19 (20.7%) | 7 (13%) | 12 (31.6%) | **0.031** |
| **Measurements at ICU admission** |  |  |  |  |
| **Temperature (°C)** | 37 (36.5; 37.6) | 37 (36.6; 37.7) | 36.8 (36; 37.2) | 0.109 |
| **Systolic pressure (mmHg)** | 120 (108.5; 130) | 120 (109; 132) | 115.6 (108; 125) | 0.274 |
| **Oxygen saturation (%)** | 92 (88; 94) | 92 (88; 94) | 91 (88; 95.5) | 0.851 |
| **Bilateral pulmonary infiltrate** | 86 (93.5%) | 51 (94.4%) | 35 (92.1%) | 0.688 |
| **Glucose (mg/dl)** | 160.5 (124.5; 208) | 152 (114; 173) | 174 (126; 267) | **0.027** |
| **Creatinine (mg/dl)** | 0.9 (0.7; 1.3) | 0.8 (0.7; 1) | 1.1 (0.7; 1.7) | **0.005** |
| **Na (mEq/L)** | 138.5 (135; 142) | 139 (136; 142) | 138 (134; 142) | 0.328 |
| **K (mEq/L)** | 3.9 (3.5; 4.4) | 3.9 (3.5; 4.4) | 4 (3.5; 4.4) | 0.763 |
| **Platelets (cell x 10<sup>3</sup>/ µl)** | 206.5 (163.5; 289.5) | 262.5 (175; 309) | 179.5 (154; 216) | **<0.001** |
| **INR** | 1.2 (1.1; 1.3) | 1.2 (1.1; 1.3) | 1.3 (1.1; 1.4) | 0.166 |
| **D Dimer (ng/ml)** | 6182 (1733; 51407) | 5403 (1940; 50079) | 6182 (976; 59829) | 0.768 |
| **LDH (UI/L)** | 489 (358; 607.5) | 502 (330; 606) | 470.4 (362; 609) | 0.962 |
| **GPT (UI/L)** | 46.5 (25.5; 69.5) | 51.5 (29; 74) | 37.5 (23.4; 69) | 0.175 |
| **Ferritin (ng/ml)** | 885 (110; 1569) | 10295 (296; 1518) | 730 (75; 1621) | 0.134 |
| **C-reactive protein (mg/dl)** | 97.6 (28; 182.8) | 74.5 (23.7; 144) | 117 (44.3; 227) | 0.211 |
| **Hematocrit (%)** | 37.7 (34.6; 40.7) | 38.4 (35.7; 41) | 35.8 (32.9; 39.6) | 0.116 |
| **WBC (cells/mm<sup>3</sup>)** | 9145 (6705; 13095) | 9450 (7400; 13720) | 8450 (6200; 11900) | 0.216 |
| **Lymphocytes (cells/mm<sup>3</sup>)** | 525 (335; 800) | 595 (400; 830) | 465 (290; 670) | 0.073 |
| **Neutrophils (cells/mm<sup>3</sup>)** | 8310 (5865; 11190) | 8395 (6630; 11480) | 7950 (5300; 10400) | 0.526 |
| **Monocytes (cells/mm<sup>3</sup>)** | 300 (170; 496.9) | 350 (200; 600) | 190 (120; 400) | **<0.001** |
| **Severity score** |  |  |  |  |
| **APACHE** | 15 (10.5; 19) | 13 (8; 16) | 18 (14; 23) | **<0.001** |
| **SOFA** | 6 (4; 8) | 5.5 (4; 7) | 7 (5; 9) | **0.013** |
| **Treatment during hospitalization** |  |  |  |  |
| **Invasive ventilation** | 88 (95.7%) | 50 (92.6%) | 38 (100%) | 0.140 |
| **Non-invasive ventilation** | 32 (34.8%) | 19 (35.2%) | 13 (34.2%) | 0.923 |
| **Hydroxychloroquine** | 91 (98.9%) | 54 (100%) | 37 (97.4%) | 0.413 |
| **Corticoids** | 79 (85.9%) | 49 (90.7%) | 30 (78.9%) | 0.111 |
| **Azithromycin** | 76 (82.6%) | 44 (81.5%) | 32 (84.2%) | 0.732 |
| **Remdesivir** | 9 (9.8%) | 7 (13%) | 2 (5.3%) | 0.298 |
| **Tocilizumab** | 29 (31.5%) | 20 (37%) | 9 (23.7%) | 0.175 |
| **Lopinavir/ritonavir** | 89 (96.7%) | 52 (96.3%) | 37 (97.4%) | 0.999 |
| **Beta Interferon** | 51 (56%) | 25 (47.2%) | 26 (68.4%) | **0.044** |
| **Time course and outcome** |  |  |  |  |
| **Hospital stay (days)** | 23.5 (17.5; 38) | 36 (21; 58) | 18 (14; 23) | **<0.001** |
| **ARDS** | 88 (95.7%) | 50 (92.6%) | 38 (100%) | 0.140 |
| **Sepsis** | 51 (55.4%) | 29 (53.7%) | 22 (57.9%) | 0.690 |
| **Septic shock** | 43 (46.7%) | 24 (44.4%) | 19 (50%) | 0.599 |

## Results

### Characteristics of the patients

Baseline characteristics of the patients according to 30-day mortality are shown in **Table 1**. Patients who died were older than those who survived (p<0.001). Besides, patients who died had more frequently arterial hypertension (p=0.012) and type-2 diabetes (p=0.031), higher values of glucose (p=0.027) and creatinine (p=0.005), lower values of platelets (p<0.001) and monocytes (p<0.001), and higher APACHE (p<0.001) and SOFA (p=0.013) scores. Eight and thirteen patients had no detectable levels in the plasma of anti-SARS-CoV-2 S IgG or anti-SARS-CoV-2 S IgM respectively. Patients who died were more frequently treated with beta interferon (p=0.044) and had a shorter hospital stay (p<0.001) than those who survived the first 30 days in ICU.

### Antibodies, antigenemia, viral RNA load and 30-day mortality

Samples from those patients with absence of anti-SARS-CoV-2 S IgM were collected at a median time since symptoms onset of 5 days, for 10 days in those with presence of IgM (*p* = 0.008). In turn, samples from those patients with absence of anti-SARS-CoV-2 S IgG were collected at a median time since symptoms onset of 8 days, for 10 days in those with presence of IgG (*p* = 0.147). Patients who died had lower frequency of positive anti-SARS-CoV-2 S IgM (**Supp file 1A**; *p* = 0.005) and IgG (**Supp file 1B**; *p* = 0.050) than those who survived, and higher frequency of plasma SARS-CoV-2 N antigenemia (**Supp file 1C**; *p* = 0.011). The Kaplan-Meier analysis showed that patients with absence of anti-SARS-CoV-2 S IgM and IgG had lower survival than patients with seropositivity of IgM (**Figure 1A**; p < 0.001) and IgG (**Figure 1B**; p = 0.003). However, patients with SARS-CoV-2 N antigenemia had lower survival than patients without N antigenemia (**Figure 1C**; p = 0.011). The Cox regression analysis adjusted by the most relevant covariates (**Table 2**) showed that the presence of anti-SARS-CoV-2 S IgM (adjusted hazard ratio (aHR) = 0.39 (95% of confidence interval (95%CI) = 0.17; 0.88); p = 0.023) and IgG (aHR = 0.17 (95%CI = 0.06; 0.44); p < 0.001) were associated with a lower risk of death. Besides, the presence of plasma N antigenemia were associated with a higher risk of death (aHR = 2.45 (95%CI = 1.27; 4.71); p = 0.007). Patients who died had lower values of anti-SARS-CoV-2 S IgM (**Supp file 2A**; p = 0.002) and IgG (**Supp file 2B**; p = 0.004) than those who survived. Conversely, patients who died had higher viral RNA load in plasma of SARS-CoV-2 N1 (**Supp file 2C**; p = 0.028) and N2 (**Supp file 2D**; p = 0.041) than survivors. Patients with low values of anti-SARS-CoV-2 S IgM and IgG had lower survival than patients with high values of IgM (**Figure 2A**; p < 0.001) and IgG (**Figure 2B**; p < 0.001) values. Conversely, patients with high plasma viral load values of SARS-CoV-2 N1 (**Figure 2C**; p < 0.001) and N2 (**Figure 2D**; p < 0.001) had lower survival than those with low values. The adjusted Cox regression analysis (**Table 2**) reported that high plasma values of anti-SARS-CoV-2 S IgM (aHR = 0.48 (95%CI = 0.24; 0.97); p = 0.040) and IgG (aHR = 0.47 (95%CI = 0.23; 0.97); p = 0.042) were associated with a lower risk of death; and high plasma viral load values of SARS-CoV-2 N1 (aHR = 2.21 (95%CI = 1.11; 4.39); p = 0.024) and N2 (aHR = 2.32 (95%CI = 1.16; 4.63); p = 0.017) were associated with a higher risk of death.

**Table 2.** Risk factors of 30-day mortality following ICU admission. Statistics: P-values were calculated by univariate Cox regression (a) and multivariate Cox regression (b) adjusted by the most relevant covariates (see statistical analysis section). Significant differences are shown in bold. Abbreviations: HR, hazard ratio; aHR, adjusted HR; 95%CI, 95% confidence interval; p-value, level of significance.

| Outcome | Unadjusted |  | Adjusted |  |
| --- | --- | --- | --- | --- |
|  | HR (95% CI) | p<sup>(a)</sup> | aHR (95% CI) | p<sup>(b)</sup> |
| **SARS-CoV-2 S IgM** |  |  |  |  |
| S IgM (+) | 0.28 (0.14; 0.59) | **0.001** | 0.39 (0.17; 0.88) | **0.023** |
| S IgM (AUC $\geq 60$) | 0.35 (0.18; 0.67) | **0.002** | 0.48 (0.24; 0.97) | **0.040** |
| **SARS-CoV-2 S1 IgG** |  |  |  |  |
| S IgG (+) | 0.31 (0.13; 0.7) | **0.005** | 0.17 (0.06; 0.44) | **<0.001** |
| S IgG (AUC $\geq 237$) | 0.31 (0.16; 0.6) | **<0.001** | 0.47 (0.23; 0.97) | **0.042** |
| **SARS-CoV-2 N-antigenemia** |  |  |  |  |
| N-antigenemia (+) | 2.1 (1.11; 3.98) | **0.022** | 2.45 (1.27; 4.71) | **0.007** |
| **SARS-CoV-2 N RNA** |  |  |  |  |
| N1 viral load ($\geq 2.156$ copies/mL) | 2.97 (1.57; 5.62) | **0.001** | 2.21 (1.11; 4.39) | **0.024** |
| N2 viral load ($\geq 3.035$ copies/mL) | 3.19 (1.68; 6.05) | **0.000** | 2.32 (1.16; 4.63) | **0.017** |

**Figure 1:**
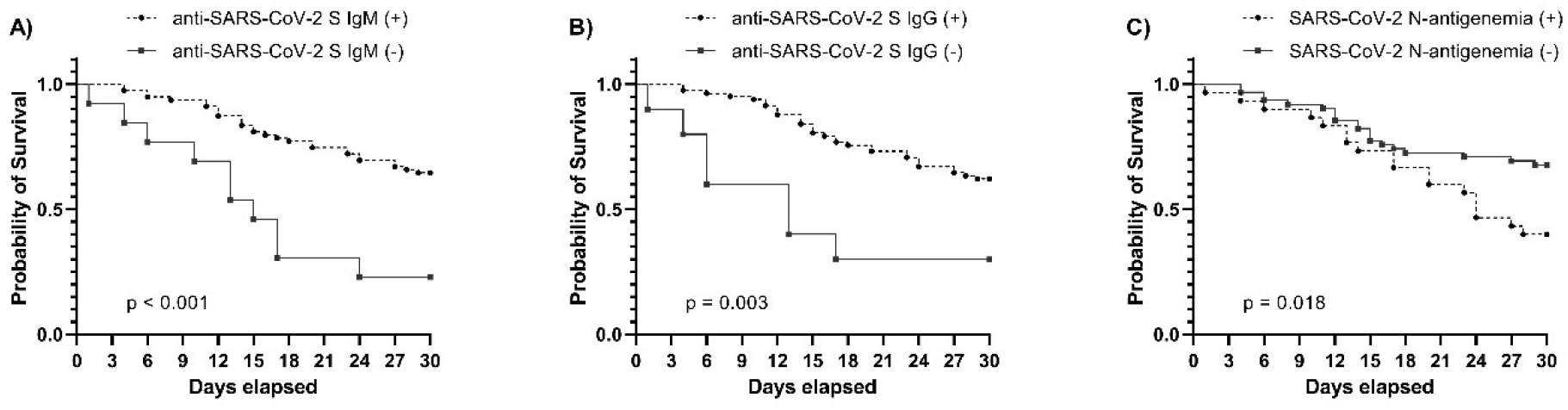
Kaplan Meier curves to represent survival by day 30 following ICU admission depending on the presence or absence of SARS-CoV-2 S antibodies (IgM, IgG) and antigenemia.

**Figure 2:**
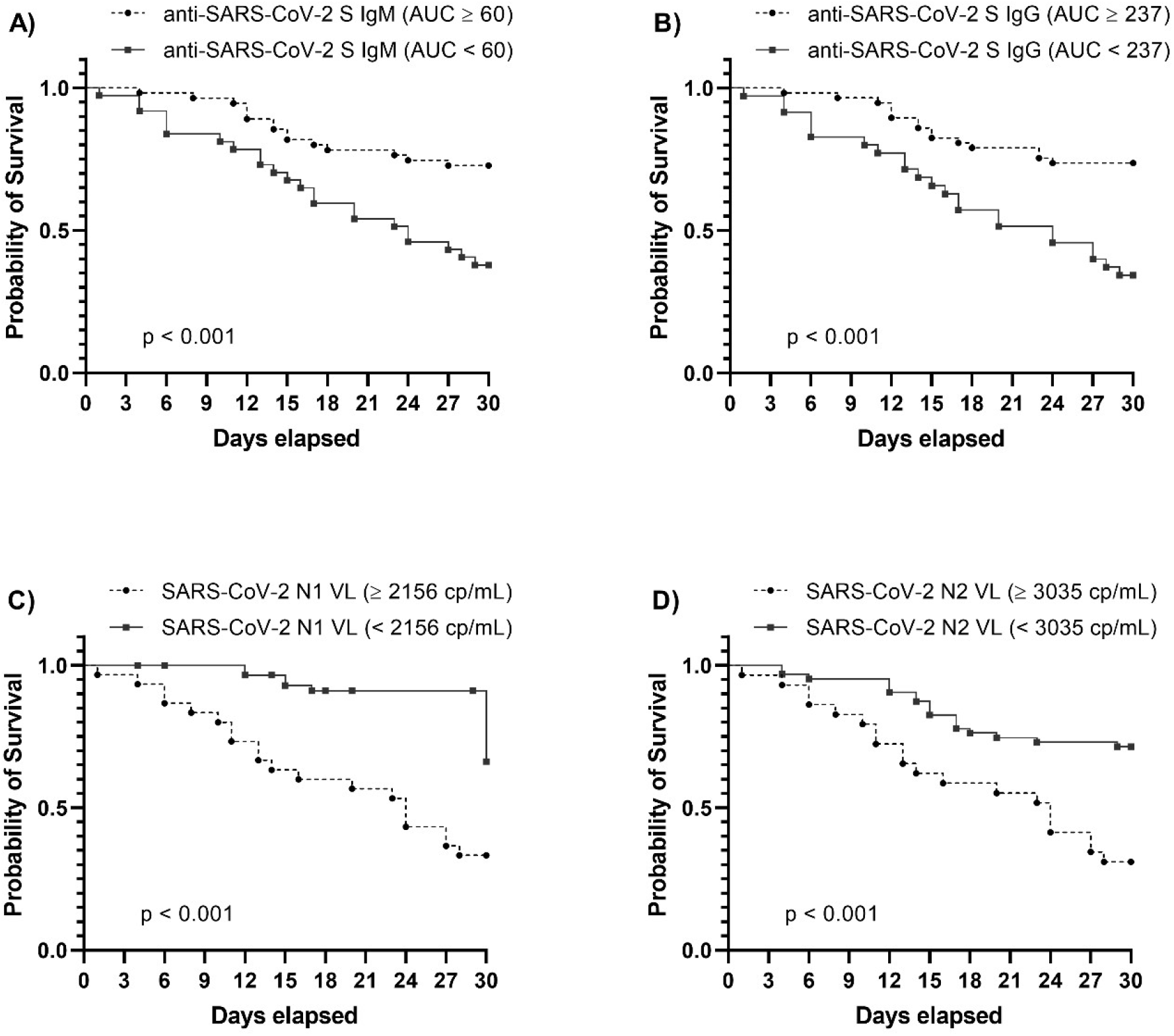
Kaplan Meier curves to represent survival by day 30 following ICU admission depending on the presence of high/low levels of SARS-CoV-2 S antibodies (IgM, IgG) or viral RNA load in plasma (N1,N2).

### Correlate between antibody responses, antigenemia and viral RNA load in plasma

Frequency of antigenemia was >2.5 fold higher in patients with absence of anti-SARS-CoV-2 S antibodies than in those who presented to the ICU with detectable antibodies (frequency of antigenemia in those patients with absence/presence of anti-SARS-CoV-2 S IgM: 77% / 25%; frequency of antigenemia in those patients with absence/presence of anti-SARS-CoV-2 S IgG: 70% / 28%) **(Figure 3)**. In turn, levels of anti-S antibodies correlated inversely with viral RNA load in plasma: (correlation coefficient, *p*): anti-SARS-CoV-2 S IgG / N1 (copies/mL) (−0.45, < 0.001); anti-SARS-CoV-2 S IgG / N2 (copies/mL) (−0.48, < 0.001); anti-SARS-CoV-2 S IgM / N1 (copies/mL) (−0.34, < 0.001); anti-SARS-CoV-2 S IgM / N2 (copies/mL) (−0.37, < 0.001) **(Figure 3)**.

**Figure 3.**
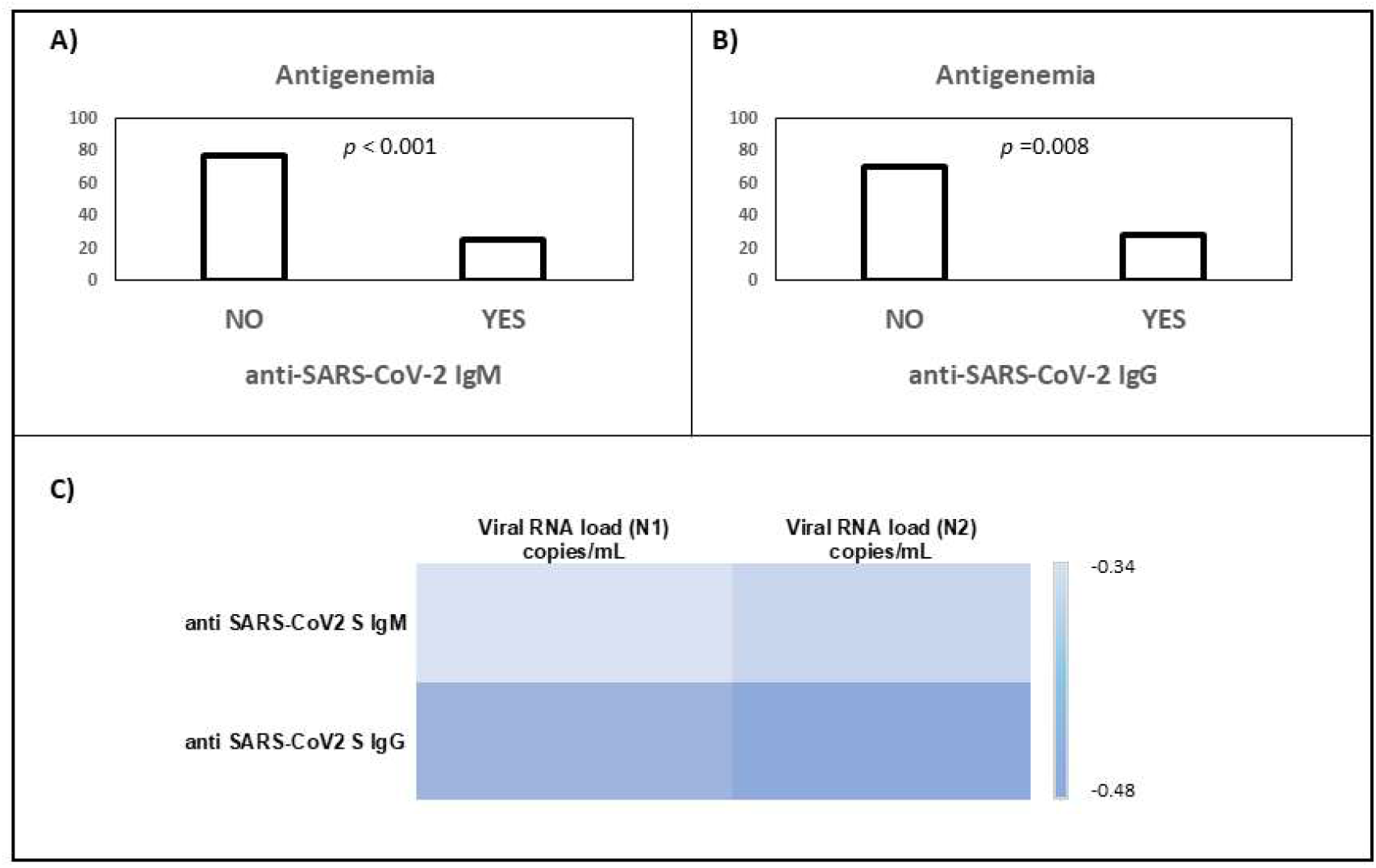
A) Frequency of antigenemia in those patients with absence or presence of anti SARS-CoV-2 S IgM. B) Frequency of antigenemia in those patients with absence or presence of anti SARS-CoV-2 S IgG. C) Heat map showing the correlation coefficients between anti SARS-CoV-2 S antibodies and viral RNA load in plasma.

## Discussion

Our study demonstrates that those critically ill COVID-19 patients with absent or insufficient levels of specific IgM or IgG antibodies against the S protein of SARS-CoV-2 following ICU admission show an increased risk of mortality. Li et al found that the production of antibodies is delayed in severe COVID-19 patients as compared to non-severe ones [10], although the former seem to exhibit higher antibody concentrations than patients with milder forms of the disease [10] [11]. The impact of antibody levels on mortality risk in COVID-19 is controversial. In the study from Röltgen K *et al*, antibody responses in acute illness were not related with patients’ outcomes [2]. In contrast, Hashem AM et al reported significantly higher levels of anti-S1 and -N IgG and IgM antibodies in patients with fatal outcomes [11]. Nonetheless, these studies include a mixture of mild, moderate and severe patients, with only a limited representation of critically ill patients. Previous work coming from our group evidence that the biological response of critically ill patients to SARS-CoV-2 infection differs from that of patients with milder forms of the disease [3]. Consequently, to study the association between levels of antibodies and mortality, it is important to analyze a homogenous population, as we do here, by considering exclusively critically ill COVID-19 patients. Results from Li K et al seem to confirm this notion. By studying only severe/critically ill COVID-19 patients, they found similar results to ours: patients non-surviving to the disease had significantly lower levels of both IgM and IgG compared to those who survived [10]. Nonetheless, in contrast to our work, the study of Li K et al lacks a multivariate analysis to confirm the association between antibody levels and mortality risk, which constitutes a major strength of our study. In turn, Asif S et al found that at both early and late timepoints following ICU admission, plasma concentrations of IgA, IgG and IgM antibodies tend to be higher in COVID-19 patients who survived compared to those who had died at 30 days, but their study involved only 19 critically ill patients [12].

Our findings suggest that the potential protective role of anti-SARS-CoV-2 S antibodies in critically patients with COVID-19 could be related to an improved control of viral replication and of dissemination of viral material at the systemic level. Patients with absent /low levels of these antibodies show higher viral RNA loads in plasma and present antigenemia more frequently, which translates into an increased risk of mortality. Although we could not determine if the presence of lower antibody levels also correlated with the presence of live virus in plasma, systemic spread of viral RNA or viral antigens can drive unspecific stimulation of the innate immune response [13], which could contribute to induce immunopathological events. Results from Li et al also support the role of antibodies in controlling SARS-CoV-2 replication, since they found higher frequencies of anti-S receptor-binding domain (RBD)-specific IgG levels in those recovered patients who were SARS CoV-2 RNA negative than those who were RNA positive in respiratory samples [10]. In agreement with our results, Röltgen K *et al* found that increases in plasma anti-SARS-CoV-2 S antibodies correlated with decreases in viral RNAemia along the course of COVID-19 [2]. In a small study with 39 patients (critically and non-critically ill), Ogata AF et al found a correlation between high concentrations of S1 antigen in plasma and ICU admission [6]. For viral-antigen positive patients, full antigen clearance in plasma was observed 5±1 days after seroconversion [6]. Recent findings in a mixed cohort also of ICU and non-ICU patients from Hingrat QL et al also suggest that clearance of antigenemia seems to be linked with the apparition of specific anti-SARS-CoV-2 antibodies [14]. As far as we know, our work is pioneer in evidencing the direct association between low antibody titers and the presence of antigenemia, and also in demonstrating the impact of antigenemia in the mortality risk of those COVID-19 patients admitted to the ICU.

In conclusion, a higher concentration of anti-SARS-CoV-2 S antibodies following ICU admission is associated with improved survival, lower antigenemia rates and lower viral RNA loads in plasma. Whether anti-SARS-CoV-2 S antibodies mediate a direct protective effect against the virus and/or reflect a broader immunological response also involving a more efficient cellular response remains to be elucidated. Further works shedding light on this regard will help to clarify the potential role of polyclonal or monoclonal antibodies against the SARS-CoV-2 S protein as treatment of critically ill COVID-19 patients [15] [16] [17]. Stratifying patients by levels of antibodies at ICU admission could help to optimize patients’ inclusion criteria, by selecting those with absence or low levels of anti-SARS-CoV-2 S IgM and IgG antibodies as those who could potentially benefit the most from these treatments. In addition, quantifying levels of anti-SARS-CoV-2 S antibodies at ICU admission could help to identify those individuals at higher risk of dying from COVID-19.

## Data Availability

Original data are available under reasonable request for scientist working in the field of COVID-19

## Availability of data and material

Original data are available under reasonable request for scientist working in the field of COVID-19

## Code availability

Not applicable

## Authors ‘contribution

SR, RA, AT, IM, JFBM, RF, FB, DJK designed the study. SR, JFBM, IM and AF drafted the manuscript. EB, LT, CA, JMG, GR, JAB, JAC, NM, PGO, RH, RC, PE, JBM, MGR, CP, FPG, MGR, JRF, SM, AM, LFB contributed with patient recruitment and data acquisition. MMV, MJMG, IM, VM, MV developed the antibody assay and profiled antibodies levels in plasma. APT, AO, NJ, JME, MDG set up the viral RNA quantification assays and profiled viral RNA load in plasma. All the authors critically reviewed the article and provided final approval of the version submitted for publication.

## Ethics approval

All procedures performed in studies involving human participants were in accordance with the ethical standards of the institutional and/or national research committee and with the 1964 Helsinki Declaration and its later amendments or comparable ethical standards. The study was approved by the Committee for Ethical Research of the coordinating institution, “Comite de Etica de la Investigacion con Medicamentos del Area de Salud de Salamanca”, code PI 2020 03 452.

## Consent to participate

Informed consent was obtained orally when clinically possible. In the remaining cases, the informed consent waiver was authorized by the Ethics committee.

## Consent for publication

not applicable

## Acknowledgements

we thank the IBSAL and CIBER administrative support to run this study.

