## Supplementary material for "Absent or insufficient anti-SARS-CoV-2 S antibodies at ICU admission are associated to higher viral loads in plasma, antigenemia and mortality in COVID-19 patients": Supp files

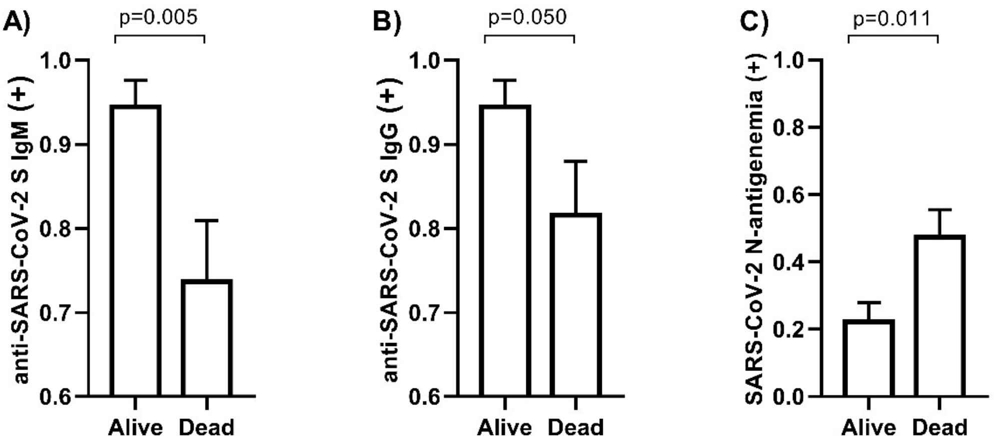


**Supplementary File 1. Frequencies of patients with positive SARS-CoV-2 S antibodies (IgM, IgG) and antigenemia by survival status 30 days following ICU admission**

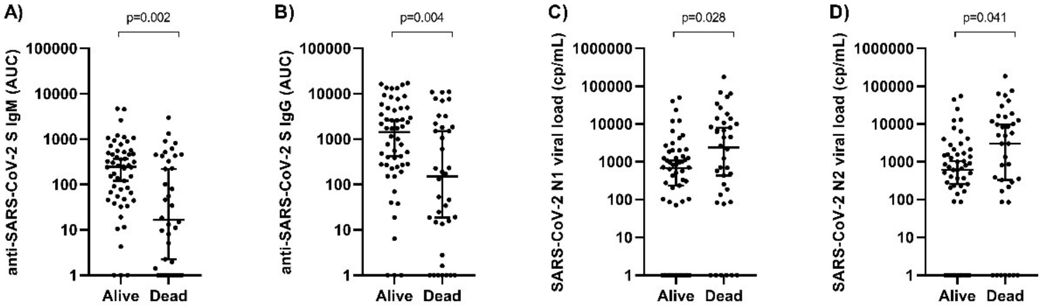


**Supplementary File 2. Levels of SARS-CoV-2 S antibodies (IgM, IgG) and viral RNA load (N1, N2) in plasma following ICU admission by survival status at day 30**
